# COVID-19-related burnout reduces COVID-19 vaccination intention in cardiac patients: a cross-sectional study in Greece

**DOI:** 10.1101/2023.01.27.23285082

**Authors:** Petros Galanis, Aglaia Katsiroumpa, Irene Vraka, Olga Siskou, Olympia Konstantakopoulou, Daphne Kaitelidou

**Affiliations:** Clinical Epidemiology Laboratory, Faculty of Nursing, National and Kapodistrian University of Athens, Greece; Department of Radiology, P & A Kyriakou Children’s Hospital, Athens, Greece; University of Piraeus, Greece; Center for Health Services Management and Evaluation, Faculty of Nursing, National and Kapodistrian University of Athens, Greece

**Keywords:** COVID-19, vaccination, intention, patients, burnout, COVID-19 burnout scale

## Abstract

**Background:** New SARS-CoV-2 variants have emerged and COVID-19 is still a public health issue, especially for vulnerable groups such as people with chronic medical conditions.

**Objective:** To investigate the impact of COVID-19-related burnout on COVID-19 vaccination intention in cardiac patients. Moreover, we investigated other possible demographic and psychological predictors of vaccination intention in cardiac patients.

**Methods:** We conducted a cross-sectional study in Greece using a convenience sample. Data collection was performed from 20 November 2022 to 10 January 2023. We measured demographic data, COVID-19-related burnout, anxiety, depression, social support, and resilience. We used the following valid tools: COVID-19 burnout scale, Patient Health Questionnaire-4, Multidimensional Scale of Perceived Social Support, and Brief Resilience Scale.

**Results:** Among patients, 45.8% were willing to accept a COVID-19 booster dose, 25.3% were hesitant, and 28.9% were unwilling. Patients experienced moderate levels of COVID-19-related burnout. After multivariable linear regression analysis, we found that increased age and decreased emotional exhaustion due to COVID-19 were associated with increased vaccination intention. Moreover, patients who have already received a booster dose had also a greater willingness to accept a new booster dose.

**Conclusions:** Identification of factors that influence patients’ decision to accept a COVID-19 booster dose is crucial to maintain a high vaccination coverage rate among them in order to avoid COVID-19-related outcomes. Since a COVID-19 booster dose on an annual basis seems to be necessary policy makers should develop and implement vaccination programmes tailored for patients.

## Introduction

COVID-19 pandemic continuous to threat public health, healthcare systems and economies causing more than 6.7 million deaths until 25 January 2023.^1^ COVID-19 patients with a known chronic medical condition have more poor outcomes (i.e., disease severity, admission to intensive care unit, invasive ventilation, and death) than COVID-19 patients without comorbidity.^2–4^ Thus, several underlying disorders, such as cardiovascular diseases, cerebrovascular disease, diabetes, hypertension, and chronic kidney disease are considered as important risk factors for COVID-19 patients.^2–4^

Several systematic reviews including a great number of studies have already shown that COVID-19 vaccines are effective and safe in patients with chronic diseases, such as malignancies, immune-mediated inflammatory disease, and immunocompromised patients.^5–9^ Moreover, evidence from studies including individuals with cardiovascular disease supports the safety and effectiveness of COVID-19 vaccines in this population group.^10,11^ That is, the Heart Failure Association of the European Society of Cardiology and the American Heart Association suggest COVID-19 vaccination as soon as possible to individuals with heart failure, heart disease, heart attack, and cardiovascular risk factors since they are high risk groups to develop COVID-19 outcomes.^12^

Although vaccination promotes public health and it is a cost-effectiveness intervention even in case of COVID-19,^13–15^ vaccine hesitancy is still one of the biggest threats to global health^16^. Socio-demographic variables, psychological factors, vaccine hesitancy, COVID-19 related variables, personal beliefs, knowledge, fear, conspiracy theories, fake news, and trust are the most important factors that influence individuals’ decision to accept the COVID-19 vaccines.^17–22^

New SARS-CoV-2 variants are emerged and COVID-19 is still a public health issue especially for vulnerable groups such as people with chronic medical conditions. Thus, vaccination uptake against COVID-19 among high-risk groups should be maintained in high levels in order to avoid the negative consequences of the disease. Therefore, the aim of our study was to investigate the impact of COVID-19-related burnout on COVID-19 vaccination intention in cardiac patients. Moreover, we investigated other possible demographic and psychological predictors of vaccination intention in cardiac patients.

## Methods

### Study design

We performed a cross-sectional study in Greece using a convenience sample of cardiac patients. Data collection was performed from 20 November 2022 to 10 January 2023. Our inclusion criteria were the following: (a) cardiac patients with a confirmed diagnosis, (b) adults over 18 years old, (c) people who can understand the Greek language, and (d) individuals that had been fully vaccinated against COVID-19 with the primary doses.

Our study was anonymous and voluntary and participants gave their informed consent. Moreover, we conducted our study according to the guidelines of the Declaration of Helsinki. Additionally, our study protocol was approved by the Ethics Committee of Faculty of Nursing, National and Kapodistrian University of Athens (reference number; 421, 10 November 2022).

### Measurements

Study questionnaire included demographic data, COVID-19-related burnout, anxiety, depression, social support and resilience. We measured the following demographic data: gender, age, educational level, self-perceived health status, SARS-CoV-2 infection, COVID-19 booster doses, and adverse effects because of COVID-19 vaccination.

We used the COVID-19 burnout scale (COVID-19-BS) to measure COVID-19-related burnout among cardiac patients.^23^ The COVID-19-BS includes 13 items and three factors (i.e., emotional exhaustion, physical exhaustion, and exhaustion due to measures against the COVID-19). Each factor takes values from 1 to 5 with higher values being indicative of higher level of COVID-19-related burnout. A previous study in a Greek sample showed that the COVID-19-BS is a reliable and valid tool.^24^ In our study, Cronbach’s alpha coefficient for the factor “emotional exhaustion” was 0.923, for the factor “physical exhaustion” was 0.856, and for the factor “exhaustion due to measures against the COVID-19” was 0.904. Thus, the reliability of the COVID-19-BS was excellent.

We measured anxiety and depression among cardiac patients with the Patient Health Questionnaire-4 (PHQ-4).^25^ In particular, we used the reliable and valid Greek version of the PHQ-4.^26^ Anxiety and depression scores range from 0 (normal levels) to 6 (severe symptomatology). In our study, Cronbach’s alpha coefficient for the anxiety was 0.871, and for the depression was 0.820 indicating very good reliability.

Social support was measured with the Greek version of the Multidimensional Scale of Perceived Social Support (MSPSS).^27,28^ Total support score ranges from 1 (low levels of support) to 7 (high levels of support). In our study, Cronbach’s alpha coefficient for the MSPSS was excellent with a value of 0.942.

Resilience was measured with the Greek version of the Brief Resilience Scale (BRS).^29,30^ Resilience score takes values from 1 (low resilience) to 5 (high resilience). In our study, reliability of the BRS was very good, i.e., Cronbach’s alpha coefficient was equal to 0.818.

We used one single item to measure patients’ intention to accept a COVID-19 vaccine. In particular, we asked patients “How likely do you think you are to get a booster COVID-19 booster dose?” and their answers were on a scale from 0 (very unlikely) to 10 (very likely). In order to categorize patients, we considered answers (a) from 0 to 2 as unwilling patients to accept a booster dose, (b) answers from 3 to 7 as hesitant patients, and (c) answers from 8 to 10 as willing patients.

### Statistical analysis

We use descriptive statistics to present categorical and continuous variables. In particular, we use numbers and percentages for the categorical variables. Also, we use mean, standard deviation, median, minimum value, and maximum value for the continuous variables. We considered demographic data, COVID-19-related burnout, anxiety, depression, social support and resilience as possible predictors of patients’ vaccination intention to accept a COVID-19 vaccine. Vaccination intention score followed normal distribution. Thus, we used univariate and multivariable linear regression analysis to assess the impact of independent variables on vaccination intention. In that case, we presented unadjusted and adjusted coefficients beta, 95% confidence intervals (CI) and p-values. Independent variables with a p-value < 0.05 in the multivariable model were considered as statistically significant. We used the IBM SPSS 21.0 (IBM Corp. Released 2012. IBM SPSS Statistics for Windows, Version 21.0. Armonk, NY: IBM Corp.) for the analysis.

## Results

Study sample included 166 cardiac patients. Mean age of the participants was 52.4 years. Most of the participants were females (77.1%), possessed a university degree (77.1%), and self-estimated their health status as very poor/poor/moderate (79.5%). Also, 61.4% of patients had been infected with SARS-CoV-2 during the pandemic.

The majority of patients have received a booster dose (91.6%). Detailed demographic data of patients are shown in Table 1.

**Table 1.** Demographic data of patients (N=166).

| Variables | N | % |
| --- | --- | --- |
| Gender |  |  |
| Males | 38 | 22.9 |
| Females | 128 | 77.1 |
| Age (years)<sup>a</sup> | 52.4 | 9.9 |
| Educational level |  |  |
| High school | 38 | 22.9 |
| University degree | 128 | 77.1 |
| Self-perceived health status |  |  |
| Very poor/poor/moderate | 132 | 79.5 |
| Good/very good | 34 | 20.5 |
| SARS-CoV-2 infection |  |  |
| No | 64 | 38.6 |
| Yes | 102 | 61.4 |
| COVID-19 booster doses |  |  |
| No | 14 | 8.4 |
| Yes | 152 | 91.6 |
| Adverse effects because of COVID-19 vaccination<sup>a</sup> | 2.7 | 2.7 |

<sup>a</sup> mean, standard deviation

Descriptive statistics for the scales in our study are shown in Table 2. Patients’ vaccination intention to accept a COVID-19 booster dose was moderate. In particular, 45.8% (n=76) of patients were willing to accept a booster dose, 25.3% (n=42) were hesitant, and 28.9% (n=48) were unwilling. Patients experienced moderate levels of COVID-19-related burnout. Also, levels of emotional exhaustion due to COVID-19 (mean score=3.29) and exhaustion due to measures against the COVID-19 (mean score=3.14) were higher than levels of physical exhaustion due to COVID-19 (mean score=2.46). Anxiety and depression symptoms were low to moderate among patients. Moreover, patients were received high levels of social support, while their resilience was moderate.

**Table 2.** Descriptive statistics for the scales in our study.

| **Score** | **Mean** | **Standard**<br>**deviation** | **Median** | **Minimum**<br>**value** | **Maximum**<br>**value** |
| --- | --- | --- | --- | --- | --- |
| Emotional exhaustion due to COVID-19 | 3.29 | 1.27 | 3.6 | 1 | 5 |
| Physical exhaustion due to COVID-19 | 2.46 | 1.12 | 2.3 | 1 | 5 |
| Exhaustion due to measures against the COVID-19 | 3.14 | 1.34 | 3.3 | 1 | 5 |
| Anxiety | 2.07 | 1.75 | 2.0 | 0 | 6 |
| Depression | 2.00 | 1.73 | 2.0 | 0 | 6 |
| Social support | 5.67 | 1.31 | 6.0 | 1.2 | 7 |
| Resilience | 3.51 | 0.78 | 3.7 | 1.8 | 5 |
| Vaccination intention | 5.64 | 3.84 | 6.0 | 0 | 10 |

Linear regression models with patients’ intention to accept a COVID-19 booster dose as the dependent variable are shown in Table 3. Univariate linear regression found that ten variables were associated with patients’ decision to accept a COVID-19 booster dose, i.e. gender, age, educational level, previous COVID-19 booster doses, adverse effects because of COVID-19 vaccination, emotional exhaustion due to COVID-19, physical exhaustion due to COVID-19, exhaustion due to measures against the COVID-19, anxiety, and depression. After multivariable linear regression analysis, we found that increased age and decreased emotional exhaustion due to COVID-19 were associated with increased vaccination intention. Moreover, patients who have already received a booster dose had also a greater willingness to accept a new booster dose.

**Table 3.**
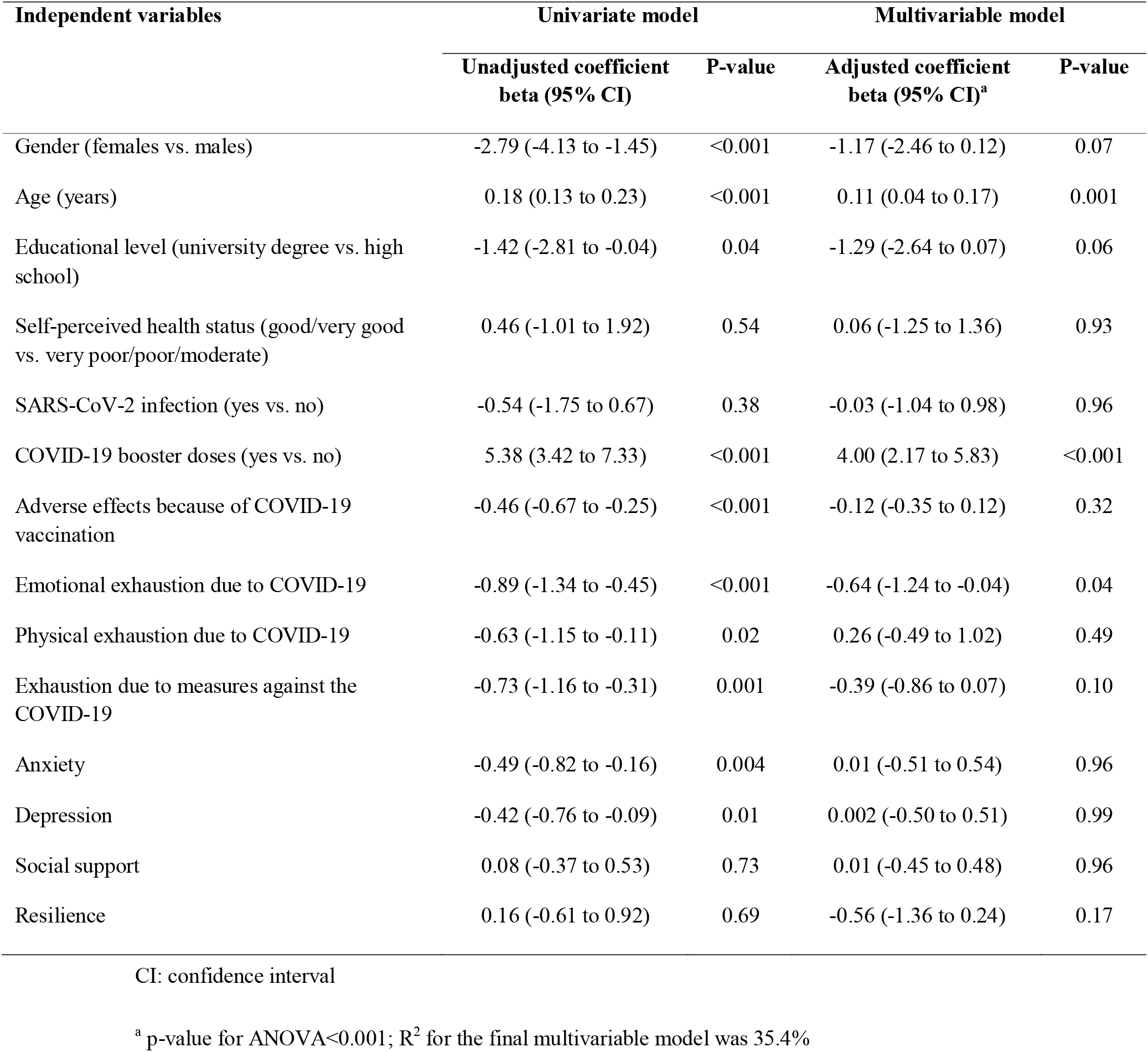
Univariate and multivariable linear regression analysis with patients’ intention to accept a COVID-19 booster dose as the dependent variable.

## Discussion

We conducted a study in Greece in order to assess the intention of cardiac patients to accept a COVID-19 booster dose. Moreover, we investigated possible predictors of such intention including demographic variables, COVID-19-related burnout, anxiety, depression, social support, and resilience.

Among cardiac patients, 45.8% were willing to accept a booster dose, 25.3% were hesitant, and 28.9% were unwilling. These results are worrying since acceptance level was low considering the fact that a meta-analysis found that 79% of the general population was willing to accept a first COVID-19 booster dose.^18^ Additionally, several studies including patients with chronic medical conditions found higher acceptance rates than our study population ranging from 54.2% to 70.4%.^31–33^ However, it is probable that these differences could be attributed to the different time that studies were conducted. Moreover, 62% of participants in a study in Greece that was conducted in May 2022 were willing to receive a second booster dose.^34^ However, later studies in Greece that were conducted during September-October 2022 found significant lower vaccination acceptance even among nurses. In particular, in a sample from the general population 34.1% reported being very likely to be vaccinated with a booster dose,^24^ while among nurses the respective percentage was slightly higher (37.1%)^35^. Moreover, low levels of seasonal influenza vaccine intention among nurses in Greece for the 2022/2023 season (57.3%) are an alarmed sign of vaccine hesitancy in the country.^36^

We found that COVID-19-related burnout affect negatively the intention of cardiac patients to accept a booster dose. Earlier studies in Greece confirm this finding including samples from the general population and nurses.^24,35^ No other studies has investigated until now this relationship throughout the world. Although we cannot make direct comparisons with other studies, we should notice that three years after the onset of the COVID-19 pandemic it is quite probable that people experience burnout. Additionally, it is well known that consequences for COVID-19 patients with comorbidity, such as disease severity, admission to intensive care unit, invasive ventilation, and death are worse than COVID-19 patients without comorbidity.^2–4^ Thus, people with a chronic disease may experience higher levels of burnout during the pandemic, as they may need to adhere to measures against the COVID-19 more than healthy people in order to avoid COVID-19-related outcomes. Moreover, emotional exhaustion due to COVID-19 may be higher among patients since they experience higher levels of fear. Literature suggests a positive relationship between fear and social and mental health outcomes during the pandemic, e.g. depression, anxiety, stress, and insomnia.^37–42^ There is a need for greater focus on patients’ mental health issues, such as COVID-19-related burnout, fear, depression, anxiety, etc.

Also, our results showed that age was associated with COVID-19 vaccination intention in cardiac patients as we expected. Literature supports this finding since several systematic reviews found that older age is a strong predictor of COVID-19 vaccination intention and uptake.^19,43,44^ Similar studies earlier in Greece suggest that increased age is associated with an increased probability of vaccine uptake.^45,46^ Several meta-analyses have shown that older age is associated with an increased risk of hospitalization, admission to intensive care unit, and death.^47–49^ Thus, older patients may feel more vulnerable against COVID-19. In addition, self-perceived risk of infection could be higher among patients. It makes sense that older patients are more afraid of the negative effects of COVID-19 and that result on a higher vaccination intention.

According to our study, patients who have received a booster dose were also more intended to accept further vaccination. A previous COVID-19 booster dose indicates the positive attitude of individuals towards vaccination. Previous studies in Greece found that seasonal influenza vaccination was related with COVID-19 vaccines intention and uptake.^34,46^ Moreover, trust in COVID-19 vaccination is a strong predictor COVID-19 vaccine uptake.^45,50^ COVID-19 vaccine hesitancy is one of the biggest threats to fighting the disease through vaccination. Several factors are the major predictors of vaccine hesitancy: socio-demographic factors, psychological factors, occupational factors, knowledge, and conspiracy beliefs.^51^ Vaccine hesitancy threatens public health for the last two decades. Concerns on safety and efficacy of vaccines, poor knowledge and information, and mistrust of public health authorities, healthcare workers, and new vaccines are significant barriers.^52–54^

Our study has several limitations. First, we used a convenience sample that cannot be representative of the cardiac patients. Second, the number of our participants was limited. Thus, further studies with bigger and random samples would add significant information. Third, we investigated several possible predictors of vaccination intention but there is still room to search for others. Fourth, we used a self-reported questionnaire and information bias could exist. Fifth, as a cross-sectional study design was used causal relationships cannot be established. Thus, there is a need for prospective studies with less bias. Finally, we measured vaccination intention that is a proxy for vaccination uptake. Therefore, future studies should measure vaccination uptake among patients in order to get more valid results.

In conclusion, we found that the percentage of cardiac patients who were willing to accept a COVID-19 booster dose was low. Moreover, we found that increased age and decreased emotional exhaustion due to COVID-19 were associated with increased vaccination intention. Identification of factors that influence patients’ decision to accept a COVID-19 booster dose is crucial to maintain a high vaccination coverage rate among them in order to avoid COVID-19-related outcomes. Since a COVID-19 booster dose on an annual basis seems to be necessary policy makers should develop and implement vaccination programmes tailored for patients. Positive attitudes towards vaccination, trust on public health authorities and elimination of conspiracy beliefs are essential to improve COVID-19 vaccination uptake.

## Data Availability

All data produced in the present study are available upon reasonable request to the authors

## Notes

### Competing Interest Statement

The authors have declared no competing interest.

### Funding Statement

This study did not receive any funding

### Author Declarations

Our study was approved by the Ethics Committee of Faculty of Nursing, National and Kapodistrian University of Athens (reference number; 421, 10 November 2022)

